# Feasibility randomised controlled trial of online group Acceptance and Commitment Therapy for Functional Cognitive Disorder (ACT4FCD)

**DOI:** 10.1101/2023.02.20.23286183

**Authors:** Norman Poole, Sarah Cope, Serena Vanzan, Aimee Duffus, Nadia Mantovani, Jared Smith, Barbara Barrett, Melanie Tokley, Martin Scicluna, Sarah Beardmore, Kati Turner, Mark Edwards, Rob Howard

## Abstract

**Introduction:** Functional cognitive disorder is seen increasingly in clinics commissioned to assess cognitive disorders. Patients complain of frequent cognitive, especially memory, failures. The diagnosis can be made clinically, and unnecessary investigations avoided. While there is some evidence that psychological treatments can be helpful, they are not routinely available. Therefore, we have developed a brief psychological intervention using the principles of Acceptance and Commitment Therapy (ACT) that can be delivered in groups and online. We are conducting a feasibility study to assess whether the intervention can be delivered within a randomised controlled trial. We aim to study the feasibility of recruitment, willingness to be randomised to intervention or control condition, adherence to the intervention, completion of outcome measures, and acceptability of treatment.

**Methods and analysis:** We aim to recruit 48 participants randomised 50:50 to either the ACT intervention and Treatment as Usual (TAU), or TAU alone. ACT will be provided to participants in the treatment arm following completion of baseline outcome measures. Completion of these outcome measures will be repeated at 8, 16, and 26 weeks. The measures will assess several domains including psychological flexibility, subjective cognitive symptoms, mood and anxiety, health related quality of life and functioning, healthcare utilisation, and satisfaction with care and participant-rated improvement. Fifteen participants will be selected for in-depth qualitative interviews about their experiences of living with FCD and of the ACT intervention.

**Ethics and dissemination:** The study received a favourable opinion from the South East Scotland Research Ethics Committee 02 on 30th September 2022 (REC reference: 22/SS/0059). HRA approval was received on 1^st^ November 2022 (IRAS 313730). The study has been registered with the ISRCTN (ISRCTN12939037). The results will be published in full in an open access journal.

**ISRCTN Registration:** https://doi.org/10.1186/ISRCTN12939037

**Grant Number:** National Institute for Health and Care Research (NIHR) grant number: NIHR202743

**Strengths and limitations of this study:**

- Novel intervention with potential to be delivered at scale
- Adherence to principles of an evidence-based intervention (ACT)
- Utilising qualitative and quantitative methodologies which is key to contextualising patient experiences in a clinically meaningful measurement framework
- Recruiting from the full range of services that assess cognition in adults in the UK
- Participants not blinded to their intervention

## Introduction

Functional Cognitive Disorder (FCD) is defined as a complaint about memory function or other cognitive process in the absence of relevant neuropathology and with evidence of inconsistency between symptoms reported, objective signs, and known phenomenology of dementia syndromes.[1] Typical complaints include forgetting an intended action while in the process of carrying it out; inability to recall well-founded memories (such as PIN numbers and names); disruptions in the flow of thoughts and conversations; word-finding difficulties; and spoonerisms. [2] While anxiety and depression are common co-morbidities, they are generally mild and do not account for the severity or nature of the symptoms reported. Patients who experience FCD tend to perform as well or a little worse on neuropsychological tests as healthy controls but better than those with mild cognitive impairment and early Alzheimer’s disease.[3] Disturbance of attention is thought to be responsible for the symptoms,[4] as with other functional neurological disorders,[5] although the underlying pathophysiological mechanisms remain unknown.

Diagnostic Memory Clinics (DMCs) are funded by clinical commissioning groups and run either by local mental health services or in partnership with acute trusts. Cognitive Neurology Clinics (CNCs) are increasingly provided by acute trusts and target younger populations with potential young onset dementias, and are more likely to see patients with FCD. As access to these services has improved, an increasing proportion of attendees are being diagnosed with FCD.[6] To date, the population prevalence of FCD has not been studied. However, a recent systematic review described 56 studies which demonstrated a high prevalence of cognitive symptoms in community populations (30% in 245,654 subjects).[3] The same review reported that 24% of DMCs attendees may have FCD, or one of its synonyms (range: 12–56%).

Those with FCD seeking referral to memory clinics have elevated levels of distress, depression, and anxiety.[3] Their symptoms persist and adversely impact employment status and activities of daily living (ADLs). Schmidtke and colleagues [7] found that cognitive symptoms persisted in 85% of those followed-up for an average of 20 months. One of their cohort (2.1%) went on to develop dementia, in keeping with the rate of revision of functional diagnoses and neurological conditions generally.[8] Hence, FCD should not be regarded as the precursor for an inevitable dementia.

It is unclear how to help this group of patients. A recent survey of DMCs found 73% immediately discharged them to primary care and treatments offered ranged from simple reassurance to referral to a community mental health team.[9] As they receive little explanation for their symptoms, patients are liable to present to their GP requesting further referrals and investigations, with the potential for iatrogenic harm and unnecessary healthcare costs.

There is now preliminary evidence that strategies focused on expectations, cognitive restructuring and psychoeducation can be helpful.[10, 11] A recent meta-analysis of treatment studies to date found that group interventions involving both cognitive-training and expectancy-modification significantly improved psychological well-being. While expectancy-change interventions had little effect on objective measures of cognitive functioning, cognitive-training was associated with small, clinically insignificant improvements in tasks related to the training ones, with no generalisation to daily life.[11] No adverse events were described.

A rare, high quality RCT included in the meta-analysis involved 18 patients receiving 13 sessions of group cognitive-behavioural therapy (CBT) intended to change participants’ beliefs and expectations.[10] Patients in the treatment group had significantly better memory-related self-efficacy than the controls at the end of the intervention and at 6-month follow-up (n=18). However, such interventions are not routinely provided by diagnostic memory, cognitive neurology, or neuropsychiatry clinics, where these patients are most likely to be seen. Furthermore, a 13-session intervention is resource intensive and unfeasible within DMCs and CNCs. Increasing Access to Psychological Therapies (IAPT) services are unlikely to offer any intervention beyond support for any co-morbid anxiety or depressive disorder.

St George’s Hospital in South London, in collaboration with South West London and St George’s NHS Mental Health Trust, is a centre of excellence for functional neurological disorders. We have developed a five-session group treatment based on the third-wave cognitive behaviour therapy known as Acceptance and Commitment Therapy (ACT). This intervention focuses on changing a person’s relationship with their thoughts and feelings. It makes use of mindfulness and acceptance processes and increases values-driven behaviour.[12] ACT considers psychological inflexibility – behaviour that is driven by attempts to excessively control internal experiences (such as difficult thoughts and feelings – as a source of emotional distress; hence, its aim is to enhance psychological flexibility.[13] Psychological flexibility is defined as “the ability to contact the present moment more fully as a conscious human being, and to change or persist in behaviour when doing so serves valued ends”[14] while psychological inflexibility is regarded a trans-diagnostic process common to numerous psychopathological states.[15]

Perceived threat is thought to be a maintenance factor for FND. Symptoms, such as cognitive failures in the case of FCD, are experienced as threatening, which causes hypervigilance and autonomic arousal. Symptoms are therefore experienced in a “top-down” manner - influenced by cognitive and neurobiological processes of expectations and predictions of illness.[5, 16] Improvement with ACT occurs through six key processes that can be grouped as either “mindfulness and acceptance processes” or “commitment and behaviour change processes”.[12] Consequently, ACT aims to change a person’s relationship with their internal experiences, increasing psychological flexibility and altering the top-down expectations by facilitating bottom-up processes, e.g., enhancing connection with direct experiences through mindfulness practices.

ACT’s efficacy across a range of conditions has been demonstrated by several RCTs and meta-analyses. A recent review of 20 meta-analyses found it to be superior to inactive controls, treatment as usual (TAU), and active interventions (excluding CBT), with effect sizes ranging from small to medium.[17] There is evidence that ACT effectively reduces distress and disability in chronic pain [18] and long-term medical conditions [19] while it has also been recommended for the treatment of functional neurological disorders generally.[13, 20] ACT can be delivered in one-to-one or group formats. It is feasible and acceptable to deliver to patients with psychosis in a brief group format [21] and can be effectively delivered online through guided and unguided modules of learning.[22]

Unpublished initial pilot data suggested improvement in measures of quality of life and decreased psychological distress in four cohorts of patients who have received our brief ACT intervention. Following this, we were able to secure funding for a feasibility study from the National Institute for Health and Care Research (NIHR; grant number NIHR202743). We now aim to study the feasibility of delivering a randomised controlled trial of ACT for FCD as an online group intervention and compare this against current treatment as usual (TAU). We also aim to further refine the ACT intervention over the course of the study so it can be adapted and manualised for a future definitive randomised controlled trial.

### Study objectives

The feasibility study aims to investigate:

- The willingness of clinicians in local services commissioned to assess patients with cognitive complaints to refer patients diagnosed with FCD into the study
- The willingness of patients with FCD to consent to the trial and be randomised to ACT+TAU vs TAU
- Acceptability of the online group ACT intervention
- Appropriateness and acceptability of clinical outcome measures
- Completion rates for outcome measures at the various time points and rate of adherence to the ACT intervention
- Fidelity of intervention
- Time needed to collect and analyse data
- Healthcare utilisation pre- and post-intervention
- Signal of efficacy in clinical outcomes

## Methods

### Trial design

ACT4FCD is a parallel-group, single-blind RCT, designed to assesses the feasibility of delivering a trial of a brief online group Acceptance and Commitment Therapy (ACT) and comparing it against the current standard intervention (Treatment As Usual (TAU)). Participants are assessed at baseline and again at 8 weeks, 16 weeks, and 26 weeks. In addition, the study aims to collect data on health utilisation before and after the intervention and has an embedded qualitative study of lived experience of FCD and the ACT intervention. This study adheres to the Standard Protocol Items Recommendations for Interventional Trials (SPIRIT).

### Recruiting sites and participants

We are recruiting participants from Diagnostic Memory Clinics (DMCs), Neuropsychiatry Clinics (NPCs) Clinical Neurology Clinic (CNCs) in London. Recruitment began on 07/11/2022. These different clinics assess a spectrum of patients with cognitive symptoms. DMCs tend to see an older population while younger patients are more often referred to the NPC and CNC. If we are not successful recruiting participants from these sites alone then we will review within the trial management group and consider adding additional recruiting sites in nearby specialist clinics.

Clinicians in the recruiting clinics will refer potentially suitable patients to the study team and deliver the “treatment as usual” (TAU) intervention as part of routine clinical care. Potential participants will provide verbal consent to being contacted by the research team. Following this, they will receive the participant information sheet (see Appendix 2), including dates of the ACT groups, and the informed consent form (see Appendix 3) and given at least 24 hours to consider these. The consent form will also record whether participants are willing to be contacted about involvement in a parallel qualitative study, described below. Potential participants will be informed that choosing not to take part will not impact their medical treatment with any service. Once consent is given, the research assistant contacts the potential participant to complete a screening interview against the inclusion/ exclusion criteria (see Figure 1. for Study Flow Chart).

**Figure 1.**
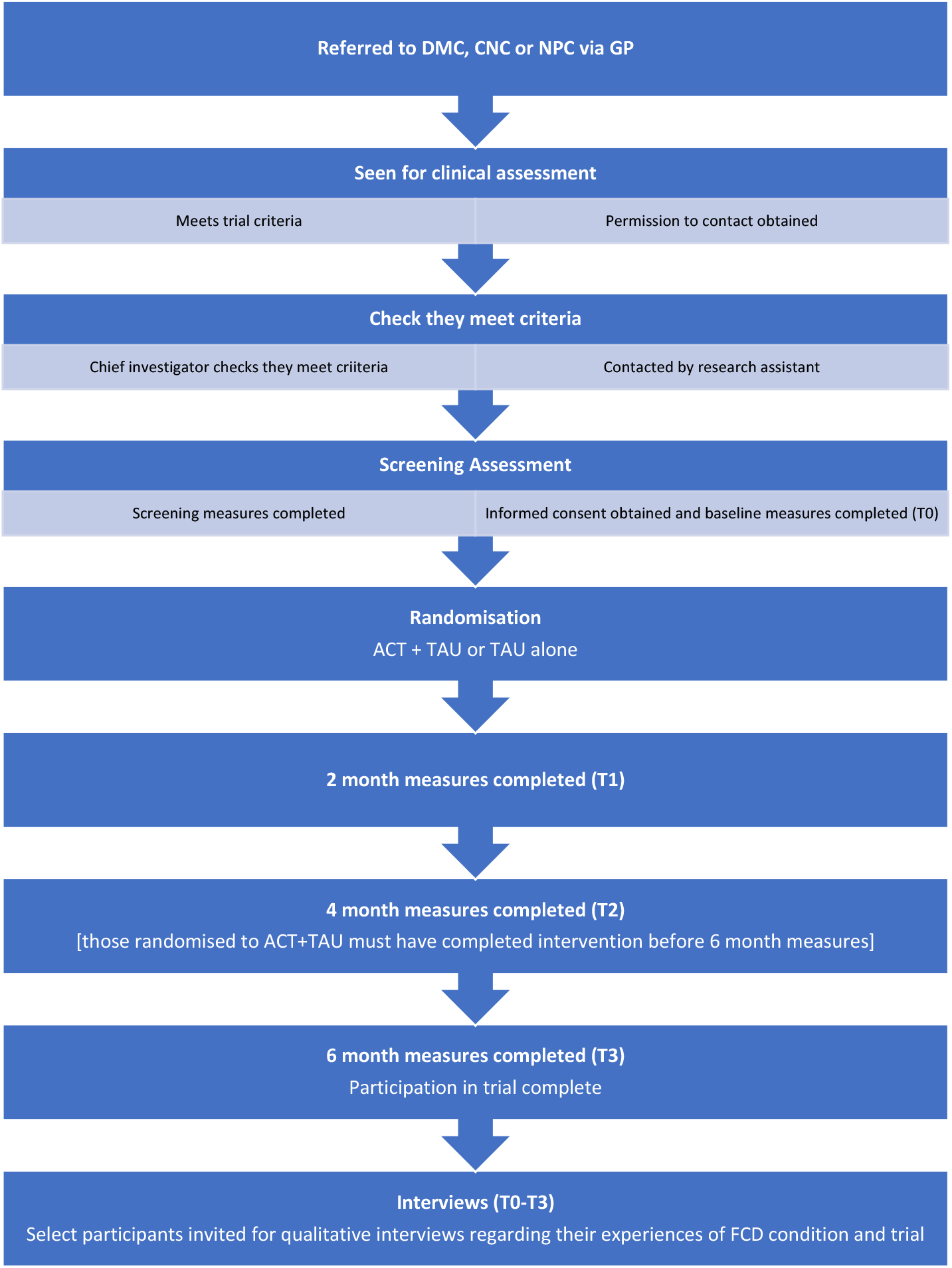
Study Flow Chart.

The inclusion/ exclusion criteria are:

#### Inclusion criteria

- An established diagnosis of FCD made in DMC/CNC/NPC and confirmed by research team from review of clinical notes and examination findings
- Aged 18 or over
- Capacity to provide written informed consent

#### Exclusion criteria

- Disabling cognitive symptoms in the context of a primary psychiatric disorder (e.g. depression, severe generalised anxiety disorder, PTSD, bipolar affective disorder, schizophrenia).
- Greater than mild-moderate depressive or anxiety disorders (PHQ9 score ≥ 15 and/or GAD7 score ≥ 15)
- At “medium” or “high” risk of deliberate self-harm and/or suicide (based on clinical assessment)
- Another predominant functional disorder (e.g., functional seizures)^*^
- Diagnosis of dementia
- Diagnosis of learning disability
- Insufficient command of English to engage in conversation without an interpreter (as this would not be compatible with the online ACT intervention)

### Primary outcome measures and progression criteria

The feasibility study is primarily gathering data on the feasibility of conducting a future definitive RCT. The primary outcome measures (and progression criteria) are:

- Rate of successful recruitment (≥ 70% intended participants recruited)
- Rate of successful adherence (≥ 75% in ACT + TAU attend four or more sessions)
- Acceptability of the ACT intervention (qualitative interview themes and majority (≥ 75%) satisfied/very satisfied on 5-point Likert scale)
- Signal of efficacy (based on increased psychological flexibility following intervention)

### Randomisation

Participants will be randomised into ACT (plus TAU) or TAU using a simple block randomisation procedure (with randomly permuted block sizes of 2 and 4). Randomisation will be carried out via the online service Sealed Envelope by the Trial Manager, who will then inform participants of their arm allocation.

### Blinding

Given the nature of the intervention, it is not possible to blind the participants to their intervention or those responsible for delivering the intervention (NP, SC, AD). The research assistant collecting the outcome data and the statistician will remain blind to treatment allocation (single-bind trial). Clinical outcome measures will be completed again at 8 weeks (T1), 16 weeks (T2), and 26 weeks (T3). Unblinding will be allowed only in case of an Serious Adverse Event (eg, resulting in death).

### Secondary (clinical) outcome measures

Those who are deemed eligible to participate in the trial will be sent an individualised weblink to complete baseline (T0) clinical outcome measures. These can be completed on paper if the participant prefers. The paper forms will be returned to the research assistant for inputting into the online database and then securely destroyed.

Clinical outcome measures will be completed at baseline (T0) and at 8 weeks (T1), 16 weeks (T2), and 26 weeks (T3). The proposed clinical outcome measures and the domains being measured include:

#### Health-related quality of life/functioning

- World Health Organisation Disability Assessment Schedule (WHODAS 2.0) [23]
- EuroQol 5-Dimension, 5-Level Health Scale (EQ-5D-5L) [24]
- ICEpop CAPability measure for Adults (ICECAP-A) [25]

#### Subjective cognitive symptoms

- Multifactorial Memory Questionnaire (MMQ) [26]

#### Depression and anxiety

- 9-question Patient Health Questionnaire (PHQ-9) [27]
- Generalized Anxiety Disorder 7 (GAD-7) [28]

#### ACT specific measure of change

- Acceptance and Action Questionnaire II (AAQ-II) [29] is a measure of psychological flexibility/inflexibility widely used in ACT. This would potentially be the primary outcome measure in a future definitive RCT.

#### Service utilisation and other cost variables

The Adult Service Use Schedule (AD-SUS) [30] is a self-report service use questionnaire completed by the study participant in interview with a trained researcher. The AD-SUS was developed by the study economist for previous work in similar populations and has been adapted for the needs of this study so that participants can complete this without assistance.

#### Improvement

- Clinical Global Impression – Improvement Scale (CGI-I), single item, participant rated

#### Measure of satisfaction

- Satisfaction rating of treatment (single-item, 5-point Likert scale)

The various research activities and outcome measures and the time points when they are collected are listed in Table 1.

**Table 1.**
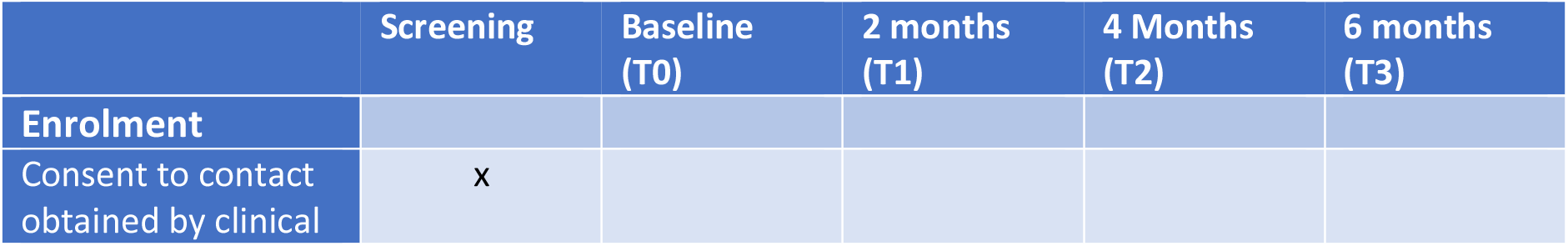
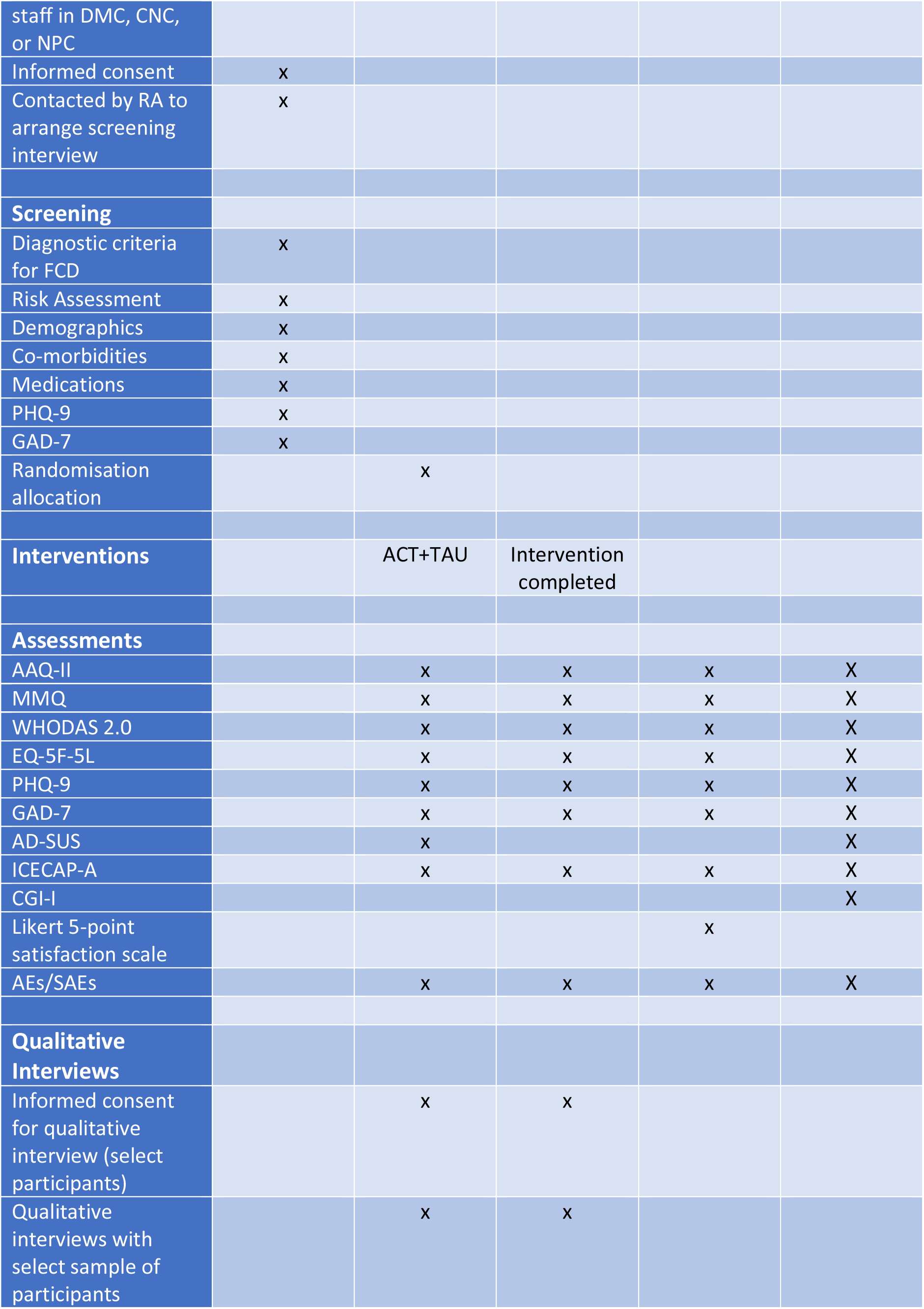
Protocol schedule of procedures for the ACT4FCD study

### Participant payment

Participants will receive a non-contingent payment of £25 for taking part in the trial. To aid retention, participants will receive £10 at each time point clinical outcome measures are fully completed. Those who take part in the optional qualitative interviews (see below) will also receive an additional £20 incentive.

### The ACT intervention

The group consists of psychoeducation about normal memory functioning, including the roles of attention and normal patterns of forgetting. The psychoeducation element aims to reduce the threat of cognitive symptoms. In line with the ACT model, the aim is to increase psychological flexibility (our proposed primary clinical outcome measure for a future RCT; Acceptance and Action Questionnaire II (AAQ-II)) in response to the symptoms and associated thoughts and feelings.

The concept of “secondary suffering” is introduced. [32] It is suggested that attempts to control cognitive “failures” leads to additional suffering, such as ruminations, negative predictions, and avoidance. It is explicitly stated that the intervention does not aim to reduce “primary suffering” (the cognitive symptoms), although it is possible that improvements *may* occur if secondary suffering is reduced. Brief mindfulness practices are incorporated in the group to facilitate “bottom up” processing, acceptance, and more neutral interpretations of unwanted experiences. Value-based goals are identified throughout the sessions in order to shift away from avoidance-based behaviour and the focus on cognitive symptoms.

Participants will be given the time of all ACT sessions prior to randomisation and are asked to consent only if able to attend should they be randomised to the active intervention. They will sent an email with link and diary invite for all the sessions the week before the first session and follow up reminder calls/emails each week to maximise attendance and engagement despite their memory problems. If a participant fails to attend a session, they will receive a follow up call from the research assistant to ensure they had the time and link and to enquire about future attendance.

The intervention protocol (see Appendix 4) will be amended in response to specific feedback received during the contemporaneous qualitative interviews which are designed to explore the participants experience of and satisfaction with the groups.

The intervention group will also receive TAU, as below.

### Treatment as usual

Treatment as usual (TAU) was selected as the fairest comparison, given that is what most patients in the UK currently receive within mental health and cognitive neurology settings. It consists of an explanation of the FCD diagnosis, provision of additional information about the diagnosis and underlying factors (such as medications, chronic pain, or poor sleep hygiene; information: https://www.neurosymptoms.org/en/symptoms/fnd-symptoms/functional-cognitive-symptoms), and signposting to local psychological services in primary care (Increasing Access to Psychological Therapies (IAPT)) for appropriate treatment, such as CBT, when co-morbid anxiety and/or depression have been identified. TAU will be delivered as part of routine clinical care by the recruiting service.

### Withdrawal and non-adherence

Participants who do not attend out of the SF-GAPI modules or assessments are not replaced. Disclosed reasons for withdrawal, non-adherence or loss to follow-up are reported.

### Sample size

This is a feasibility trial; as such, a power calculation is neither possible nor necessary. Rather, the sample size is pragmatic. Target recruitment is 48 patients in total (24 in each treatment arm), which provides sufficiently reliable estimates of feasibility outcomes such as recruitment, adherence and retention rates to inform a fully powered RCT. For example, assuming 70% of those approached consent to participate in the trial, the 95% confidence interval for adherence rate would have width of 13.0%. For those in the treatment arm, the 95% confidence interval for an intervention adherence rate of 80% would have width of 16.0%. A sample size of 48 is also consistent with those recommended for pilot and feasibility studies to provide adequate data and precision of means and variances (*n* between 24 and 50). [33, 34, 35] If it proves challenging to recruit participants via these routes alone then we will approach colleagues at other specialist services in London who see patients with FCD to increase our recruitment pool.

### Statistical analysis plan

A fully documented statistical analysis plan, centred on describing key process measures to decide if a definitive trial is feasible, will be prepared by the statistician (SAP), formally agreed with co-investigators and the Trial Steering Committee prior to data collection being completed. Participant throughput will be summarized in an extended Consolidated Standards of Reporting Trials (CONSORT) diagram.[36] The CONSORT flow chart will be used to present descriptive data on trial referral, enrolment, intervention allocation, adherence, and retention and to document any deviations from protocol.

Feasibility outcomes will be summarised using descriptive statistics, with 95% confidence intervals (CIs) provided to permit assumptions when planning a future definitive trial. Data relating to referral, screening and enrolment, and recruitment logs will be used to produce accurate estimates of eligibility, consent, and recruitment rates.

Treatment assignation, intervention adherence (e.g., ACT session attendance) and satisfaction of care data will be used to contribute to the evaluation of the acceptability of randomisation and allocated intervention/treatment arms. At each time point, the time taken (per participant) to complete trial measures will be recorded. Retention rates will also be estimated for each of the patient-reported/clinical outcome measures, with consideration given to differential dropout between the arms of the trial to identify potential (attrition) bias in treatment completion and/or data collection. All feasibility outcomes will be compared with full-trial progression criteria.

Baseline characteristics will be reported according to treatment arm. Continuous variables will be reported as mean (standard deviation (SD)) if normally distributed or median (inter-quartile range) if non-normal, while categorical variables will be presented as frequency (%). Subsequent analyses will summarise the proposed patient-reported and clinical outcomes (e.g., psychological flexibility, subjective memory function, quality of life, and depression/anxiety measures) at each time point for each trial arm using appropriate descriptive statistics (e.g., group mean, SD). To provide an indication of potential changes in scores/frequencies between the four time points, linear/logistic mixed effects regression models will be employed performed on an intention-to-treat (ITT) basis (accounting for data assumed to be missing at random). These random intercept (mixed) models will include intervention group, time, and intervention group by time interaction. There will be no emphasis on hypothesis testing, however, which is reserved for the future main trial. Rather, pre-to-post-intervention standardised effect sizes (Hedges’ g, relative risk) will be computed (SDs will be computed from estimated model standard errors) with associated CIs calculated to explore imprecision around effect sizes.[37] Due to the small sample size, important covariates (e.g., baseline score on relevant measure, gender, age) may be included in models if the two arms happen to be highly imbalanced. ITT analyses will also be administered by imputing values for missing data using a conservative last observation carried forward (LOCF) procedure (given full sensitivity analysis testing of missing data assumptions is beyond the scope of a feasibility study). Exploratory analyses using mixed effect models will examine the rate of change in intervention and control groups on outcome measures across four time-points, adjusting for relevant baseline scores and variables of interest (anxiety, depression, subjective evaluation of memory), and investigate changes on a per-protocol basis (focused on intervention adherence; i.e., including only participants who attended at least 4 sessions and with post-treatment data).

### The qualitative study

A sub-sample of participants in both arms of the trial will be invited for in-depth semi-structured interviews over the course of the intervention period. A sampling framework will be used that ensures participants are included that are representative of the sociodemographic characteristics and clinical profile. Interviews at baseline (T0) will focus on the experience of living with FCD and subsequent interviews (at T1), not necessarily with the same participants, will explore their views on the ACT intervention. These interviews will focus on the acceptability, number, and frequency of ACT sessions, ways of optimising engagement, perceived benefits/limitations of the intervention, and any recommendations for improvement of the components and/or content of the intervention.

Interview schedules will be coproduced by the PPI representatives and the research team. One-to-one, in-depth interviews will be carried out via Teams by the research assistant, recorded, and transcribed, and the transcripts cross-checked against the original recordings to ensure accuracy. The analysis will be led by the qualitative study expert and research assistant using reflexive thematic analysis [38] aided by NVivo12 software. The results of the T1 interviews will be utilised to optimise the intervention during the feasibility study with the aim of developing a formalised intervention to be used in a future RCT.

### Data management

All data will be inputted by the participants themselves on to the trial database on REDCap and backed up weekly on a secure server. The electronic Trial Master File will be backed up weekly on an additional encrypted hard drive. No paper copies will be stored. Please refer to Data Management Plan (see Appendix 5) for details regarding confidentiality, data collection, data handling, and data transfer. The data collection and management will be in line with GDPR Data Protection Act (2018) and SWLSTG’s Information Governance.

## Data Availability

All data produced in the present study are available upon reasonable request to the authors

## Patient and public involvement

The research design has been informed by our PPI representatives who were recruited from earlier pilots of the intervention conducted within the SWLSTG Neuropsychiatry Service. They assisted in the development of the intervention, study methodology, and review of clinical outcome measures. Two PPI participants (SB, MS) have been recruited to the Trial Steering Committee and are funded to assist with reviewing study materials, writing, and developing patient information leaflets, producing the semi-structured interview schedules for the qualitative study, and ensuring proper conduct of the study. We will document PPI activity over the course of the study in order to accurately assess where and how the lived experience perspective has been used and its impact on the research process and findings. All PPI representatives have lived experience of FCD and the proposed ACT intervention.

## Serious adverse events

All adverse events (AEs) and serious adverse events (SAEs) reported spontaneously by participants or observed by researchers will be recorded and reported to the Trail Manager (TM) and Chief Investigator (CI). Urgent actions concerning participant and staff safety, communication with others, and clinical care will be immediately addressed by the CI and reported to the Trial Management Group (TMG). A summary of (S)AEs will be presented at each Trial Management Group (TMG) and Trial Steering Committee (TSC) meeting. AEs will be categorised for severity and seriousness by the TM and CI. SAEs will be further reviewed for relatedness to trial procedures and unexpectedness by the CI initially, and additionally by the chair of the TSC.

## Ethics and dissemination

Ethics approval was sought from the South East Scotland Research Ethics Committee (REC) 02 and a favourable opinion was received on 30^th^ September 2022 (REC reference: 22/SS/0059). Any amendments to the protocol will be agreed with the REC before being implemented and then amended on the ISCTRN Registry. The findings of the study will be published on an open access journal once the full trial has been completed. A data monitoring committee was not deemed necessary as participants are adverse effects are not expected in either of the randomisation groups.

## Author contributions

NP, SC, and ME conceived the study. NP, SC, ME, NM, JS, and RH, developed and finalised the study design. JS provided statistical expertise in the clinical trial design and NM provided qualitative research expertise in the qualitative study design. BB contributed to the design of the health economics analysis. SB, MS, and KT provided expert by experience expertise. All authors contributed to refinement of the study protocol. NP drafted the manuscript. All authors provided critical revisions to the manuscript and approved the final manuscript. NP is grant holder. NP, SC, SV, JS, ME, RH, and BB will have access to the final trial dataset. The authors intend to share deidentified individual clinical trial participant-level data after publication of results with researchers who provide a methodologically sound proposal. Requests can be sent to

## Study sponsor details

South West London & St George’s Mental Health NHS Trust (SWLSTG)

Sponsor contact:

Tania West

R&D Governance & Clinical Trials Manager

South West London & St George’s Mental Health NHS Trust

Research & Development, Newton Building (1st Floor)

Springfield University Hospital

Glenburnie Road

London SW17 7DJ

## Sponsor roles and responsibilities

SWLSTG Mental Health NHS Trust is not involved in any aspect of the study design and management, or in its data collection, analysis or interpretation. The Sponsor is responsible for providing the research team with adequate arrangements to successfully complete the trial, and to monitor its compliance with ethical guidelines and legislation. SWLSTG Mental Health NHS Trust is the data controller for the study and all data shall return to the Sponsor at the end of the trial. A collaboration agreement is being finalised with King’s College London, where a Co-Investigator and a Trial Statistician are based. No Data Sharing Agreements are required.

## Funder roles and responsibilities

NIHR is not involved in any aspect of the study design and management, or in its data collection, analysis or interpretation. Core publications (including, but not limited to, research protocol and results) were agreed with NIHR before the beginning of the trial, and NIHR will be informed of any additional publications. NIHR will also be acknowledged in any public dissemination of the research, including peer-reviewed articles, conference presentations and patient involvement activities.

## Trial steering committee roles and responsibilities

The TSC will oversee the study on behalf of the of the trial Sponsor and Funder and ensure that the study is conducted within appropriate NHS and professional ethical guidelines. It will provide advice on all appropriate aspects of the project; will oversee progress of the trial, adherence to the protocol, participant safety and the consideration of new information of relevance to the research question; will ensure the rights, safety and well-being of the participants are given the most important considerations; will ensure appropriate ethical and other approvals are obtained in line with the project plan; will agree proposals for substantial protocol amendments and provide advice to the Sponsor and Funder regarding approvals of such amendments. It will comprise the CI (NP), TM (SV), independent Chair (Professor Michael Kopelman), independent member (Dr Laure McWhirter), sponsor representative, and two independent PPI members.

## Trial management group roles and responsibilities

The TMG is composed of AD, BB, JS, ME, NM, NP, RH, SB, SC, SV. The agreed terms of reference are:

1. To review and provide feedback on any amendment of the study protocol, outcome measures and recruitment documents
2. To support the progress of the trial towards its interim and overall objectives and against the study timescale
3. To agree on a trial monitoring plan
4. To advise on participant recruitment and retention
5. To facilitate data analysis
6. To help troubleshoot challenges to the timely completion of the trial
7. To review the study outcomes
8. To discuss future developments of the study
9. To report to the Trial Steering Committee (TSC)

## Data availability statement

The data that support the findings of this study will be openly available in ISRCTN Registry at https://doi.org/10.1186/ISRCTN12939037

## Footnotes

* Co-morbid functional diagnosis is acceptable so long as those symptoms do not dominate the clinical picture.

