## Appendix 2 for "Feasibility randomised controlled trial of online group Acceptance and Commitment Therapy for Functional Cognitive Disorder (ACT4FCD)"

***Chief Investigator: Dr Norman Poole***

### Participant Information Sheet

We'd like to invite you to take part in our research study. Before you decide, it is important that you understand why the research is being done and what it would involve for you. Please take time to read this information and discuss it with others if you wish. If there is anything that is not clear, or if you would like more information, please contact the research team (see details at the bottom of this leaflet).

**What is the aim of this study?**

People with Functional Cognitive Disorder (FCD) suffer from memory problems that negatively impact their everyday life and personal wellbeing. At present treatment options are limited in number and accessibility. Acceptance and Commitment Therapy (ACT) focuses on changing a person's relationship with their thoughts and feelings, using mindfulness and acceptance. ACT has been successfully used in the treatment of conditions similar to FCD, and we have tested ACT on FCD in one small study where it appeared to work well.

Now we need to try it with a larger number of patients from different clinics and different backgrounds to make sure it is acceptable and useful to all patients. When we introduce a new way of working it is important that we compare it against our usual way of working and measure the results to see which way is best. To do this we will be allocating half (50%) of all people who take part into the usual care treatment group and half (50%) into the usual care plus specialist ACT group.

To try and make sure the groups are the same to start with, each participant is put into a group by chance (randomly). This process is called randomisation and to ensure that it is fair, the group you will be allocated to will be decided by a computer programme in a clinical trials unit. None of the researchers or members of the care team will have any input into which group you will be allocated to. The findings will help us to further develop specialist 'ACT' support programmes. This is a feasibility study (a practice-run before doing a large-scale study). It will help us find out more about:

a. whether people with functional cognitive disorder find this intervention helpful; and how we can improve it.
b. whether people who have functional cognitive disorder find this type of trial acceptable and whether they are willing to be randomly allocated to receive the specialist support we have developed, in addition to their usual care; or receive only the usual care that is currently available to them.
c. how best to train staff to deliver this specialist support.
d. how best to measure the costs of that support and how well it works.
e. whether people taking part can complete the questionnaires we plan to use, without difficulty

Once we have completed the study and gathered information about each of these points, we can then make changes to prepare for conducting a larger study. We need to ensure that research meets patient needs and are asking your help to do this. If you are interested in taking part, please read the rest of this information sheet.

**Why have I been invited?**
We are asking you to take part because you have an established diagnosis of functional cognitive disorder that resulted in using one of the outpatient clinics and you are aged 18 or over. Your consultant has helped us to identify who to ask. We are inviting a total of 48 participants like you to take part.

**Do I have to take part?**
No. It is up to you to decide whether or not to take part. If you do decide to take part you will be
given this information sheet to keep and be asked to sign a consent form. If you decide to take part you are still free to withdraw at any time and without giving a reason. This would not affect your legal rights.

**What will happen to me if I take part?**
After you have consented, the research assistant will conduct an initial assessment to confirm you are eligible to take part in the study. If found to be eligible, you will be asked to complete several questionnaires assessing your quality of life, mood, memory, and how well you are functioning with the condition.

After this initial assessment the trial manager will contact King's College clinical trials unit (an external provider) who will allocate (randomise) you into one of the two groups, either the ACT intervention or standard medical care. The trial manager will then contact you to let you know which group you have been allocated to.

Whichever arm of the trial you are allocated to, you will be asked to complete questionnaires on 4 separate occasions; at baseline, and again 2 months, 4 months and finally at 6 months after first being randomised. After completing them on this 4$^{th}$ and final occasion you will leave the study.

The questionnaires allow us to study whether the ACT intervention has been helpful and whether any improvements are maintained beyond the end of the intervention. A member of the research team will be on hand to assist with completing the questionnaires, should this be required. Each set of questionnaires will take between 35 - 45 minutes to complete.

**What are the interventions I could receive?**

You will receive either:

a. the usual care and access to services provided for people following a diagnosis of functional cognitive disorder within this Trust (consisting of the standard care already provided to you in the local memory clinics, neuropsychiatry service, or cognitive neurology clinic (wherever you were seen for assessment and diagnosis).
b. the usual care and access to services provided for people following functional cognitive disorder within this Trust **plus** the specialist ACT intervention being tested in this study.

There is a possibility that you may be disappointed by which group you have been allocated to, but each of the groups **is equally important** to developing this specialist ACT support programme and we hope that whatever the outcome you will continue to take part.

<u>During the 6 months in the study</u>

If you are allocated to the usual care group you will receive all the usual support and access to services provided by your care team.

If you are allocated to receive the specialist ACT programme from the clinician, you will be contacted by the treating clinician to arrange for the group intervention which consists of 4, 2-hour weekly treatment sessions, followed by 1, 2-hour booster session a month after the final treatment session. The ACT Group Intervention will host roughly 5-8 participants per group and will be entirely online. You will be expected to attend all 5 sessions if allocated to this group. Over the study period we will run groups on different days of the week and times of the day to maximise the chance of there being a group convenient to you. However, once allocated a particular group you will remain in that one for the duration of your intervention. They are run online to minimise disruption to your schedule.

The ACT intervention includes education about normal memory and forgetting. The intervention aims to decrease the threat of memory failures and to alter a person's relationship to their symptoms while encouraging behaviour that is in line with their values and goals. The concepts of "primary suffering" (the unwanted memory symptoms) and "secondary suffering" (attempts to control memory failures) are discussed. Brief mindfulness practices are incorporated into the sessions to improve acceptance, aiming for less distressing interpretations of unwanted experiences. Value-based goals are identified throughout the sessions to shift away from avoidance-based behaviour and the focus on memory symptoms.

<u>What happens during a session?</u>

All the sessions are run by two of the researchers who have expertise in delivering online group psychological therapies. In each of the sessions, specific topics are covered, and exercises undertaken. Each session begins with an overview of what will be covered and a reminder of what had been discussed at the previous one. Members of the group are encouraged to participate openly in these sessions and share their thoughts and experiences. Homework is set at the end of each session and discussed at the subsequent one. At the end of each session some tasks are given for trying out before the next session. Feedback is then shared at the next session. Participants are actively encouraged to engage in sessions as this appears to strengthen positive outcomes of the treatment. Everything discussed is confidential so must not be shared outside the group, and this will be explained again at the start of each session.

<u>Recording of Sessions</u>

All sessions of the ACT Intervention will be video recorded using MS Teams. The purpose of recording is not to review participants, but to ensure that the ACT process has been followed. The recordings will be reviewed for ACT fidelity and deleted at the end of the trial. Participants have the right to switch off their camera during sessions if they do not wish to be recorded.

<u>Follow-up</u>

While in the study (regardless of which group you are in) we would like to follow your progress and will ask you to complete questionnaires (either online or paper copies can be sent to you) at the **start** of the study and again at **2**, **4** and **6** months. When these questionnaires are due the research assistant will contact you to confirm that you are happy to continue and arrange a mutually convenient time to meet with you in person or online to support you in completing the questionnaires. These questionnaires assess how you are doing so are very important for the study.

We will also collect information from you and your medical notes about the amount of support you have received previously and which services you have accessed. We will only collect information directly relevant to your participation in this study and nothing else.

Additionally, as part of a sub-study, we may wish to interview you for about 45-60 minutes face-to-face, either via an online videocall or in person. This interview will be audio recorded and then transcribed. Like all of your data, the recording and transcription will be anonymised. We will ask you about the support you have received and the things you found useful or most helped you in your day-to-day functions. If you are invited to attend the interview, this will be a one-off occurrence and entirely voluntary, and you will be given further information beforehand so that you can decide whether you would like to be interviewed or not. **You do not have to agree to this interview to be able to take part in the study.**

**Travel expenses and payment for participation**

Participants will receive £25 for taking part in the trial, plus reimbursement of any travel costs involved, and £10 for completing each full set of questionnaires. We will also pay £20 to those who take part in the additional 45-60 minutes interview.

**What are the possible disadvantages and risks of taking part?**

We do not expect there are any disadvantages or risks to you. We will arrange any interviews at times to suit you. You may feel anxious before or tired after taking part in the sessions or while completing the questionnaires, but we will do everything we can to minimise or prevent this. You will be asked about your well-being over the course of the study, and you will have the opportunity to take short rest breaks when completing the questionnaires.

You may find the ACT sessions tiring or inconvenient, however, the timing of these will be arranged to be maximally convenient to participants. We will run groups on two days of the week at different times so if one group runs at a time that is inconvenient then you will have the option of an alternative day/time.

**What are the possible benefits of taking part?**

We cannot promise that being in either arm of this trial will help you but the information we obtain will help us plan a larger study to test how effective the ACT intervention is. In the future, this could help improve services for other people who have a functional cognitive disorder.

**What happens when the study stops?**
When the study ends after 6 months, you will continue with your usual care from your hospital or GP.

**What will happen if I don't want to carry on with the study?**
Your participation is voluntary, and you are free to withdraw at any time, without giving any reason, and without your legal rights being affected. If you withdraw then the information collected so far cannot be erased and this information may still be used in the project analysis.

**What if there is a problem?**
If you have a concern about any aspect of this study, you should ask to speak to the Principal Investigator Dr Norman Poole or Trial Manager Dr Serena Vanzan (contact details below) who will do their best to answer your questions.

If you remain unhappy and wish to complain formally, you can do this through the NHS Complaints Procedure. Details can be obtained from the Trust's Patient Advice and Liaison Service (PALS). Tel: 0203 513 6150 (Monday - Friday 9.30am to 4.30pm) or.

**How will we use information about you?**
We will need to use information from your medical records for this research project. This information will include your:
- NHS/Hospital number
- Name
- Contact details
- Ethnicity
- Gender
- DoB
- Relationship status
- Employment status
- Current medication
- Medical history

People will use this information to do the research or to check your records to make sure that the research is being done correctly. People who do not need to know who you are will not be able to see your name or contact details. Your data will have a code number instead. We will keep all information about you safe and secure.
Once we have finished the study, we will keep some of the data so we can check the results. We will write our reports in a way that no-one can work out that you took part in the study.

**What are your choices about how your information is used?**

You can stop being part of the study at any time, without giving a reason, but we will keep information about you that we already have.

- We need to manage your records in specific ways for the research to be reliable. This means that we won't be able to let you see or change the data we hold about you.
- If you agree to take part in this study, you will have the option to take part in future research using your data saved from this study. If you consent to this we may:
    - Use your data already collected for this study in future research.
    - Contact you regarding taking part in future research relating to this current study.

**Where can you find out more about how your information is used?**

You can find out more about how we use your information

- at www.hra.nhs.uk/information-about-patients/
- by asking one of the research team
- by ringing us on 020 8725 3786

**Will my taking part in this study be kept confidential?**

We will follow ethical and legal practice and all information about you will be handled in confidence.

If you join the study, some parts of your medical records and the data collected for the study will be looked at by authorised persons from South West London Mental Health Trust, who are organising the research, and the King's Clinical Trials Unit. It may also be looked at by authorised people to check that the study is being carried out correctly. All will have a duty of confidentiality to you as a research participant and we will do our best to meet this duty.

All information which is collected about you during the course of the research will be kept strictly confidential, stored in a secure and locked office, and on a password protected database. Any information about you which leaves the hospital will have your name and address removed (anonymised) and a unique code will be used so that you cannot be recognised from it.

Your personal data (address, telephone number) will be kept for 6 months after the end of the study so that we are able to contact you about the findings of the study and possible follow-up studies (unless you advise us that you do not wish to be contacted). All other data (research data) will likewise be kept securely for 5 years after completion of the whole study (not just your involvement in it). After this time your data will be disposed of securely. During this time all precautions will be taken by all those involved to maintain your confidentiality, only members of the research team will have access to your personal data.

Although the information we collect about you is confidential, should you disclose anything to us which we feel puts you or anyone else at risk, we may feel it necessary to report this to the appropriate persons.
We would ask for your permission for the anonymised data set to be used to inform future projects and for education purposes and this permission is included within your consent form.

**Involvement of the General Practitioner (GP)**
If you do decide to take part in the study, we will inform your GP and provide them with a copy of this information sheet.

**What will happen to the results of the research study?**
We will use these findings to support the design of a large-scale study to test whether this ACT intervention results in people with functional cognitive disorder having better quality lives. The findings will be written up and submitted for publication to enable other NHS services to learn from our experiences. All reports and publications will be anonymised, and you would not be identified in any report or publication.

**Who is organising and funding the research?**
The study is being organised by the South West London and St Georges Mental Health NHS Foundation Trust (SWLSTG) and is being funded by National Institute for Health and Care Research (NIHR) Programme (project number NIHR202743).

**Who has reviewed the study?**
All research in the NHS is looked at by independent groups of people, called a Research Ethics Committee to protect your interests. This study has been reviewed and received a favourable ethical opinion from South East Scotland REC 02 (22-SS-0059).

**Further information and contact details**
If you have any questions about the study, wish to discuss taking part or have any concerns, you can contact the researchers leading the study:

| | | | |
|---|---|---|---|
| Chief Investigator | Dr Norman Poole | | 07519668140 |
| Trial Manager | Dr Serena Vanzan | | |
| Research Assistant | Aimee Duffus | | |

General information about taking part in research studies can also be obtained from your hospital's Patient Advice and Liaison Service (PALS), Tel: 0203 513 6150 (Monday - Friday 9.30am to 4.30pm) or.

Many thanks for reading this information sheet. Please keep this information sheet. We will ask you to sign a consent form if you agree to take part and we will give you a copy of it to keep.
