## Appendix 3 for "Feasibility randomised controlled trial of online group Acceptance and Commitment Therapy for Functional Cognitive Disorder (ACT4FCD)"

***Chief Investigator: Dr Norman Poole***

### Informed Consent Form

Participant Number:______________________

**Please <u>initial</u> box**

| | |
|---|---|
| 1. I confirm that I have read the information sheet dated 16.11.22 (version 2.1) for this study. I have had the opportunity to consider the information, ask questions and have had these answered satisfactorily. | ☐ |
| 2. I understand that my participation is voluntary and that I am free to withdraw at any time without giving any reason, without my medical care or legal rights being affected. | ☐ |
| 3. I understand that the data collected up to the point of my withdrawal will still be used for analysis. | ☐ |
| 4. If randomly allocated to the ACT intervention group, I consent to be video recorded during therapy session. | ☐ |
| 5. I understand that relevant sections of my medical notes and data collected during the study, may be looked at by individuals from the research team, where it is relevant to my taking part in this research. I give permission for these individuals to have access to my records. | ☐ |
| 6. I understand my personal data will be kept for a maximum of 6 months following the end of the study and anonymised data will be made available to the research team to support future research. | ☐ |
| 7. I agree to my General Practitioner being informed of my participation in the study. | ☐ |

8. I agree to take part in this study. ☐

9. I agree to be contacted to take part in an additional interview with the research team to share my experiences of living with FCD and of taking part in this trial. I understand that agreeing to be contacted does not oblige me to participate in the additional interview. Yes ☐ No ☐

10. I agree to be contacted to take part in future research regarding treatment for FCD. I understand that agreeing to be contacted does not oblige me to participate in any further studies. Yes ☐ No ☐

11. I would like to receive the study outcomes at the end of the study. Yes ☐ No ☐

| | | |
|---|---|---|
| **Name of Participant** | **Date** | **Signature** |
| Name of Person taking consent | Date | Signature |
