## Appendix 4 for "Feasibility randomised controlled trial of online group Acceptance and Commitment Therapy for Functional Cognitive Disorder (ACT4FCD)"

### Summary of ACT for FCD Group Intervention

The group includes psychoeducation on normal memory functioning, in particular the role of attention and normal patterns of forgetting. This psychoeducation element aims to reduce the threat of cognitive symptoms. In line with the ACT model, the group aim is to increase psychological flexibility in response to the symptoms and associated thoughts and feelings. ACT is a contextual cognitive-behavioural therapy approach. Instead of targeting specific thoughts in relation to unwanted symptoms, it focuses on altering a person's relationship with their symptoms, increasing acceptance of the symptoms, and encouraging behaviour that is in line with a person's values (moving away from behaviour that focuses on avoidance of symptoms).

The concepts of "primary suffering" (the unwanted cognitive symptoms) and "secondary suffering" (attempts to control cognitive "failures" causes additional suffering, such as ruminations, negative predictions, and avoidance) are introduced. It is explicitly stated that the intervention does not aim to reduce "primary suffering" (cognitive symptoms), although it is possible that improvements may occur if secondary suffering is reduced. Brief mindfulness practices are incorporated in the group to facilitate "bottom up" processing, acceptance, and more neutral interpretations of unwanted experiences. Value-based goals are identified throughout the sessions to shift away from avoidance-based behaviour and the focus on cognitive symptoms.

The intervention protocol will be amended in light of specific feedback received during the contemporaneous qualitative interviews which are designed to explore the participants experience of and satisfaction with the groups.

#### Summary of the session content

| | Group content |
|---|---|
| **Session 1** | Ground rules/house-keeping<br>Introductions<br>Psychoeducation (models of memory, normal forgetting, attention, functional cognitive disorder, fight/flight response)<br>Primary and secondary suffering<br>Vicious cycles in FCD<br>Brief mindfulness practice<br><br>Home practice:<br>Read Session 1 handout and write down any questions |
| **Session 2** | Brief mindfulness practice |

| | |
|---|---|
| | Check-in regarding home practice<br>What is ACT<br>Mindfulness<br>Values<br><br>Home-practice:<br>Mindfulness practice<br>Read Session 2 handout and write down any questions |
| **Session 3** | Brief mindfulness practice<br>Check-in regarding home practice<br>Value-based action<br>Value-based goals (passengers on the bus metaphor)<br><br>Home practice:<br>Working towards value-based goal<br>Mindfulness practice<br>Read Session 3 handout and write down any questions |
| **Session 4** | Brief mindfulness practice<br>Check-in regarding home practice<br>Cognitive fusion: Thoughts as barriers<br>Complete Willingness & Action plan<br><br>Home practice:<br>Working towards value-based goal<br>Mindfulness practice<br>Read Session 4 handout and write down any questions |
| **Session 5 (booster)** | Review goals<br>Trouble-shooting |

Summary of ACT for FCD Group Intervention

The group includes psychoeducation on normal memory functioning, in particular the role of attention and normal patterns of forgetting. This psychoeducation element aims to reduce the threat of cognitive symptoms. In line with the ACT model, the group aim is to increase psychological flexibility in response to the symptoms and associated thoughts and feelings. ACT is a contextual cognitive-behavioural therapy approach. Instead of targeting specific thoughts in relation to unwanted symptoms, it focuses on altering a person's relationship with their symptoms, increasing acceptance of the symptoms, and encouraging behaviour that is in line with a person's values (moving away from behaviour that focuses on avoidance of symptoms).

The concepts of "primary suffering" (the unwanted cognitive symptoms) and "secondary suffering" (attempts to control cognitive "failures" causes additional suffering, such as ruminations, negative predictions, and avoidance) are introduced.

It is explicitly stated that the intervention does not aim to reduce “primary suffering” (cognitive symptoms), although it is possible that improvements may occur if secondary suffering is reduced. Brief mindfulness practices are incorporated in the group to facilitate “bottom up” processing, acceptance, and more neutral interpretations of unwanted experiences. Value-based goals are identified throughout the sessions to shift away from avoidance-based behaviour and the focus on cognitive symptoms.

The intervention protocol will be amended in light of specific feedback received during the contemporaneous qualitative interviews which are designed to explore the participants experience of and satisfaction with the groups.
