## Appendix 5 for "Feasibility randomised controlled trial of online group Acceptance and Commitment Therapy for Functional Cognitive Disorder (ACT4FCD)"

### Data Management Plan (DMP)

#### POON1002

##### 1. Trial information

| | |
|---|---|
| Trial type | Non-CTIPM Intervention - Feasibility |
| Study Sites | Single-site (SWLSTG) |
| Total sample size | 48 |
| Total duration of study<br><br>(months) | Trial start date _01/05/2022__<br><br>Planned duration for recruitment (months)___6____<br><br>Planned duration of follow-up (months)___6____<br><br>Total Duration ___24____Months |
| Study objective and design | We aim to study the feasibility of delivering a Randomised Control Trial (RCT) of an online group intervention based on Acceptance and Commitment Therapy (ACT) adapted for those with FCD. The treatment aims to reduce the threat of the memory failures and improve quality of life despite their presence. Participants will be recruited via SWLSTG and St George's neuropsychiatry services and memory clinics. All those eligible will be randomly allocated to either 5 sessions of online group ACT for FCD or treatment as usual. Questionnaires measuring various dimensions of physical, cognitive and mental wellbeing will be administered 2, 4 and 6 months after initial baseline. We will compare the outcome of those who receive the intervention with treatment as usual. |

| | |
|---|---|
| | A selection of participants will be invited to take part in interviews about their lived experiences of FCD as well as to receive feedback on the study intervention. |
| Primary outcome measure | Increased psychological flexibility |
| Secondary outcome measures | Self-evaluations (questionnaires) of:<br>• Memory<br>• Anxiety<br>• Depression<br>• Global functioning<br>• Quality of life<br>• General wellbeing<br>• Service utilisation |
| Interim analysis | Yes/**no**, if yes when_____________(Months from baseline) |

#### 2. Trial personnel and contact details

*This section details the name, their position in the trial, email address, telephone/fax number for all staff involved in the trial including the sponsor. The trial coordinator/trial manager, the investigators, study staff involved in the data management (including computing staff responsibilities for maintaining hardware and software), the monitors and anyone else associated with the trial at each site.*

*2.1 Sponsor site personnel (add or remove accordingly)*

| Role | Name | Organisation | Contact Details |
|---|---|---|---|
| Sponsor | SWLSTG R&D | SWLSTG | |
| Chief Investigator | Norman Poole | SWLSTG | |
| Trial Manager | Serena Vanzan | SWLSTG | |
| Research Assistant | Aimee Duffus | SWLSTG | |
| Research Assistant | Rebecca Cox | SWLSTG | |
| Research Assistant | Tasnim Fakira | SWLSTG | |
| Co-Investigator and Trial statistician | Jared Smith | SGUL | |
| Co-Investigator and Trial statistician | Nadia Mantovani | SGUL | |
| Co-Investigator | Sarah Cope | SWLSTG | |

| | | | |
|---|---|---|---|
| Co-Investigator | Mark Edwards | KCL | |
| Trial statistician | Barbara Barrett | KCL | |
| Primary contact for DM issues | Serena Vanzan | SWLSTG | |
| Secondary contact for DM issues | Norman Poole | SWLSTG | |

#### 3. Milestones

##### *3.1 Study Milestones*

| Milestone | Date/Estimated Date |
|---|---|
| Date funding confirmed | 23.07.2021 |
| Date and version number of approved protocol | 14.07.2022, v1.0 |
| Date and version number of final protocol amendment(s) | n/a |
| Date/version number of final approved CRF | No approval required |
| Release date/version number of final database | Est end Oct 2022 |
| Date DMP signed off | |
| Date of first participant first visit (FPFV) | Est 17.10.2022 |
| Date last participant last visit (LPLV) | Est Sep 2023 |

##### *3.2 Proposed Data Milestones*

| Milestone | Date/Estimated Date |
|---|---|
| Data entry to commence | n/a (data collected directly on database) |
| Date of interim data partial lock if applicable | n/a |
| Data entry completed | Est Sept 2023 |
| Last query resolved in the system | Est Oct 2023 |
| Date of database final lock | Est Oct 2023 |

#### 4. Data collection & data entry system

*4.1* *Detail how data will be collected and entered from each site, whether to* ***complete the paper CRFs*** *or how to enter data electronically from each site.*
CRFs and outcome measures (questionnaires) will be completed electronically via the software REDCap, either by the RA (CRFs) or the participant (questionnaires). Where the participant will prefer paper questionnaires, these will be provided and returned via post, and the RA will enter the responses onto REDCap.
In the event that the online database will not be finalised at the time of recruitment opening, paper CRF and Questionnaires will be completed by the RA and participants, and the data will be entered onto REDCap as soon as this will become available.
Interview data (audio) will be automatically transcribed into a Word file by Teams, and the files will be saved in the study folder on SWLSTG's shared drive.

*4.2* *Provide details on the system used for data entry*
The REDCap online software will be used for collecting CRF and quantitative data.

*4.3* *Outline who will be conducting the data entry from each site?*
The RAs and the participants themselves.

4.4 *Outline whether single or double data entry will be carried out for all sites*
Single.

4.5 *If double data entry is required describe the process and whether a 100% of CRFs or a sample of CRFs will be double entered. Outline how the two entries will be compared, who will carry out the comparison and how the results will be dealt with.*
N/a

4.6 *Outline the checks undertaken for outcome measures*
The TM will perform regular self-monitoring visits in which the data entered for a proportion of participants will be checked for completeness, accuracy and timely completion.

4.7 *Provide details on how the data will be centralised*
N/a – one site.

#### 5. Data checks & data validation for each site

5.1 *Outline who will perform data cleaning, missing data checks, consistency checks, range checks and logic, and how these are checked at each site?*
The TM will perform regular self-monitoring visits in which the data entered for a proportion of participants will be checked for completeness, accuracy and timely completion. In addition, value range restrictions will be applied in REDCap so that no values outside the available range can be entered when completing closed questions.

5.2 *Describe how regularly the data will be checked*
Monthly.

5.3 *Provide contact person for each site for data queries if data managed centrally*
N/a

5.4 *Describe the data flow from each site to central data centre, and who will conduct the overall data check*
N/a – one site.

5.5 *Detail the **flow of the data** from the field to the final storage for each site*
Quantitative data will be collected directly on study database. Qualitative data (interviews) will be recorded on Teams (on a Trust laptop), transcribed by Teams and saved on the study's shared drive folder.

#### 6. AE and SAE data handling

*Outline how the AE data will be collected by each site and collated for all sites at the end of the trial.*
N/a – one site.

#### 7. Partial and/or final data check and database lock

*Detail if an interim analysis is planned in the trial protocol, stating the time point and whether the database will be partially locked for the interim analysis.*
N/a

*Detail the process for partial and final data checks and data lock, outline who checks the data and who will sign off for partial/final data lock form(s)?*
The database will be locked after the following actions have been confirmed as completed by the TM:

1. All CRF data has been collected and entered onto REDCap.
2. All queries identified during regular data checks and self-monitoring visits have been resolved/clarified.
3. All missing information has been confirmed as being not available (as opposed to not entered).
4. All monitoring visits (incl. close-out) have been performed, and outstanding actions completed.
5. A final data quality check has been performed.

The TM will be responsible for locking the database (notifying the CI and Sponsor), send the relevant data to the trial statisticians (with treatment allocation coded to prevent unblinding, where relevant), and save a copy of the full database in the TMF.

#### 8. Data security and transmission between sites

*Provide details on the data security procedures for transmitting data between sites*
To comply with SWLSTG's Information Governance regulations, the datasets will be sent via encrypted email to the trial statisticians.

#### 9. Data export & analysis

*Explain how data will be exported and who the data should be sent to for data analysis.*
The qualitative data will be saved on Word files, sent via an encrypted email to Nadia Mantovani, Trial Statistician and Co-Investigator. The full database with demographic and questionnaire data will be exported in CSV format and sent via encrypted email to Jared Smith, Trial Statistician and Co-Investigator. The responses to the AD-SUS questionnaire will be exported in Excel format and sent via encrypted email to Barbara Barrett, Trial Statistician.

#### 10. Data Back-up and archiving

*Describe procedures in place to ensure data protection including back-up system (if you don't do this you could lose the data!)*
The database (all data on REDCap) will be downloaded and backed up weekly on a Trust external hard drive. The TMF, including the interview transcriptions, will be saved on the Trust's shared drive, which is backed up every night, in a restricted folder dedicated to the study. In addition, the TMF will also be backed up once a week on the Trust encrypted external hard drive.
